# Scaling haplospecific antisense oligonucleotides from N-of-1 to broad use in genetic disease populations by diplotyping

**DOI:** 10.64898/2026.01.28.26345012

**Authors:** Olivia Kim-McManus, Pagé Goddard, Shahad Olsson, Liana Protopsaltis, Joseph Gleeson, Qing Zhang, Nafeesa Kahn, Ali Crawford, Stephen F. Kingsmore

## Abstract

Antisense oligonucleotides (ASO) are versatile disease modifying therapies for genetic diseases. An accelerated FD) pathway enables ASO treatment trial initiation in single patients within a year. However, this rapid N-of-1 pathway lacks extensibility to broad use necessary for sustainability. Individualized ASOs bind pre-mRNAs encompassing an entire locus. Thus, ASOs targeting common heterozygous polymorphisms (SNPs) are potentially haplospecific in many patients with dominant disorders. We developed haplospecific ASOs for two patients with *SCN2A*-Complex Neurodevelopmental Disorder (CND) and gain-of-function (GOF) or mixed gain-loss dysfunction (GLD) variants. The ASOs targeted reference *SCN2A* intronic sequences containing SNPs. The patients each had *SCN2A* haplotypes with reference SNP alleles *in cis* with causal variants and alternate SNP alleles *in cis* with normal mRNA. Following N-of-1 demonstration of safety and efficacy, we evaluated their applicability to 21 *SCN2A*-CND patients using whole genome sequencing (WGS) with haplotyping by read proximity. Ten (48%) patients had ASO-eligible diplotypes. Haplotype analysis of 1000 Genomes Project (1kGP) participants revealed 16 additional *SCN2A* haplotypes present in >20% of subjects, tagged by 156 SNPs. *In silico* assessment of specificity and potency identified additional haplospecific ASOs for validation in reprogrammed 1kGP cells heterozygous for the tagging SNPs. A combination of 4 haplospecific ASOs provided coverage for 76% of 1kGP subjects, potentially scaling N-of-1 FDA applications in future *SCN2A*-CND patients with GOF/GLD variants by ASO selection by diagnosis by WGS with haplotyping. Thus, population resources have potential to prepare haplospecific ASO therapies *a priori* for many patients and genetic diseases, with individual selection by WGS haplotyping.

**One Sentence Summary:** Population genome sequencing with haplotyping identifies haplospecific antisense oligonucleotides for disease modifying therapy of genetic disorders.

## INTRODUCTION

A triumph of medical research has been the discovery of the molecular basis of over 10,000 genetic diseases. Most genetic disorders, however, currently lack disease modifying therapies, despite orphan drug incentives and the recent advent of genetic therapy platforms. There are two main reasons for this: Firstly, for most genetic disorders either their molecular basis (such as the dysfunctional gene) or their molecular mechanism of disease (such as specific gain-of-function, GOF) was only recently discovered. Secondly, genetic diseases are rare and, as yet, the methods for their routine identification – such as diagnostic WGS – are not widely used by physicians. The resultant lack of identified affected individuals is a major impediment to perceived return on investment for pharmaceutical development. Remarkably, this Aristotelian vacuum has started to be filled by individual clinicians working with individual affected families on N-of-1 therapy trials.

The practice of medicine has always included trials of interventions in individual patients, such as a trial of labor or trial of a specific treatment. Such N-of-1 trials were historically highly attractive to individual clinicians and patients since the results applied directly to that patient.^1^ N-of-1 trials have had two prior rounds of popularity. The first, about sixty years ago, formalized experimental designs for N-of-1 research trials^2–4^. A patient would receive a therapy of interest during one time period and a placebo or alternative therapy in another period. A prespecified outcome was measured during each period. The use case was to examine the utility of an approved intervention in an individual patient with a disorder for which either no randomized controlled trial of effectiveness had been conducted, or where the individual or his/her disorder differed materially from those in such trials. These represented early examples of patient-centered research, wherein all trial aspects were negotiated by the physician and patient together.^1^ As practicable, early N-of-1 trials were blinded, randomized regarding the order of the two treatments, and had single or multiple crossovers. Initially, N-of-1 trials were limited to individual patients and lacked ability to generate broader evidence of effectiveness. Subsequently, however, some N-of-1 trials with similar patients, treatments, and outcome measures were aggregated to attempt to generate general evidence.^5,6^ Despite their potential to accelerate pharmaceutical innovation, however, N-of-1 aggregation has not become mainstream.

Interest in N-of-1 trials was resurgent 15 years ago as part of the then new paradigm of precision (personalized) medicine.^7–10^ The principle underlying precision medicine was that prevention and treatment are more effective and less likely to suffer adverse effects if tailored to measurable individual or subgroup differences than a “one-size-fits-all” approach. While stratification of treatment in patient subgroups is now becoming standard of care, bespoke N-of-1 practice has not become common.

In the last six years, N-of-1 trials reached new prominence when used in breakthrough genetic therapies for individual patients with ultra-rare genetic disorders.^11–13^ These N-of-1 trials are quite different from prior iterations for three reasons: Firstly, the targets of these therapies are hyper-rare causal variants in individual ultra-rare disorders. Thus, an N-of-1 trial is, by definition, the only method whereby they can be evaluated. Secondly, these genetic therapies are first-in-class, disease modifying interventions for severe, childhood-onset diseases without effective therapies. They make use of relatively new genetic therapy platforms, such as antisense oligo nucleotides (ASO) or Clustered Regularly Interspaced Short Palindromic Repeats (CRISPR) and CRISPR-associated (Cas) proteins. ASOs and CRISPRs are considered therapeutic platforms since many aspects of development, while unique to a molecular target, are shared across many targets^14^. ASOs, for example, have proven effective as therapies for several genetic diseases and to date thirteen have been approved by the FDA or European Medicines Agency.^15–17^ Thirdly, they have benefited from unique FDA provisions: For well-evidenced genetic therapy platforms, an accelerated process enables specific therapy development of a for an unique molecular target and a single individual treatment trial in as little as a year.^18–20^ For eligibility, such individualized ASOs must be developed for causal variants that are only present in one or two patients affected by severe or rapidly progressive genetic diseases lacking standard therapies. This process has allowed more than 40 individuals to receive individualized ASO therapies on a not-for-profit, philanthropic or research grant basis.^21,22^ Given such success, the question now is how to scale N-of-1 trials to sustainable therapies for rare disease communities^14,23,24^.

Here we describe an N-of-1 to N-of-many ASO therapy pathway based on gene-specific haplotyping. We reasoned that N-of-1 trials have substantial value beyond individual patients by accrual of evidence of safety and effectiveness for a genetic therapy platform, specific disorder, disease mechanism, delivery route, dose intensity, and set of outcome measures. At the outset of new drug development, each of these items is a critical unknown^24^. We hypothesized that N-of-1 ASOs may be an efficient starting point for drug approval for genetic diseases, acting to de-risk and accelerate the process whilst immediately benefiting individual patients.

## RESULTS

### N-of-1 use of an ASO for SCN2A-Complex Neurodevelopmental Disorder

We developed an ASO (nL-SCN2A-002, n-Lorem, San Diego) specifically for a teenage boy with a pathogenic, heterozygous variant in the voltage-gated Na^+^ channel α2 subunit (*SCN2A* c.2558G>A, p.R853Q, ClinVar ID:194555) (Kim-McManus et al, submitted). He had daily seizures that were refractory to antiepileptics, severe global neurodevelopmental delay, was non-verbal and non-ambulatory. He also had Autism Spectrum Disorder (ASD) with irritability, agitation, ataxia and choreoathetosis. These represented dominantly inherited *SCN2A*-Complex Neurodevelopmental Disorder (*SCN2A*-CND, MedGen ID:1800189) and encompassed Developmental and Epileptic Encephalopathy 11 (MIM:613721) and ASD/intellectual disability (ID)^25^. The voltage-gated Na^+^ channel NaV1.2 helps initiate and conduct neuronal action potentials^26^. The prevalence of *SCN2A*-CND is 8.9/100,000 births.^25^ p.R853Q has been reported in over 20 children with *SCN2A*-CND and has been shown to confer complex, mixed gain/loss dysfunction on neuronal Na^+^ channels.^27–32^

nL-SCN2A-002 was a 20 nucleotide (nt) phosphorothioate 2’methoxyethyl gapmer. It targeted reference sequences within *SCN2A* intron 15 on chromosome 2 (NC-000002.12: g.165322220-165322241=) located 20 kb from the patients causal *SCN2A*-CND variant. ASO specificity was due to the patient being heterozygous for rs72874313 (g.165322227A>G), a common single nucleotide polymorphism (SNP) located within the ASO target region. Furthermore, the two rs72874313 alleles were constituents in two heterozygous chromosome 2 haplotypes. Haplotype one was g.[165322227=; 165342465G>A], corresponding to rs72874313 reference *in cis* with the causal *SCN2A* variant c.2558G>A. Haplotype two was g.[165322227A>G; 165342465=], corresponding to rs72874313 alternate allele *in cis* with an *SCN2A* reference coding domain (Fig. S1). Thus, nL-SCN2A-002 bound and degraded *SCN2A* pre-mRNA with haplotype one preferentially to *SCN2A* haplotype two, which encoded a normal SCN2A protein. Upon FDA accelerated authorization and IRB approval, nL-SCN2A-002 was administered at 60 – 90 day intervals (https://Clinicaltrials.gov NCT06314490).

**Fig. S1:**
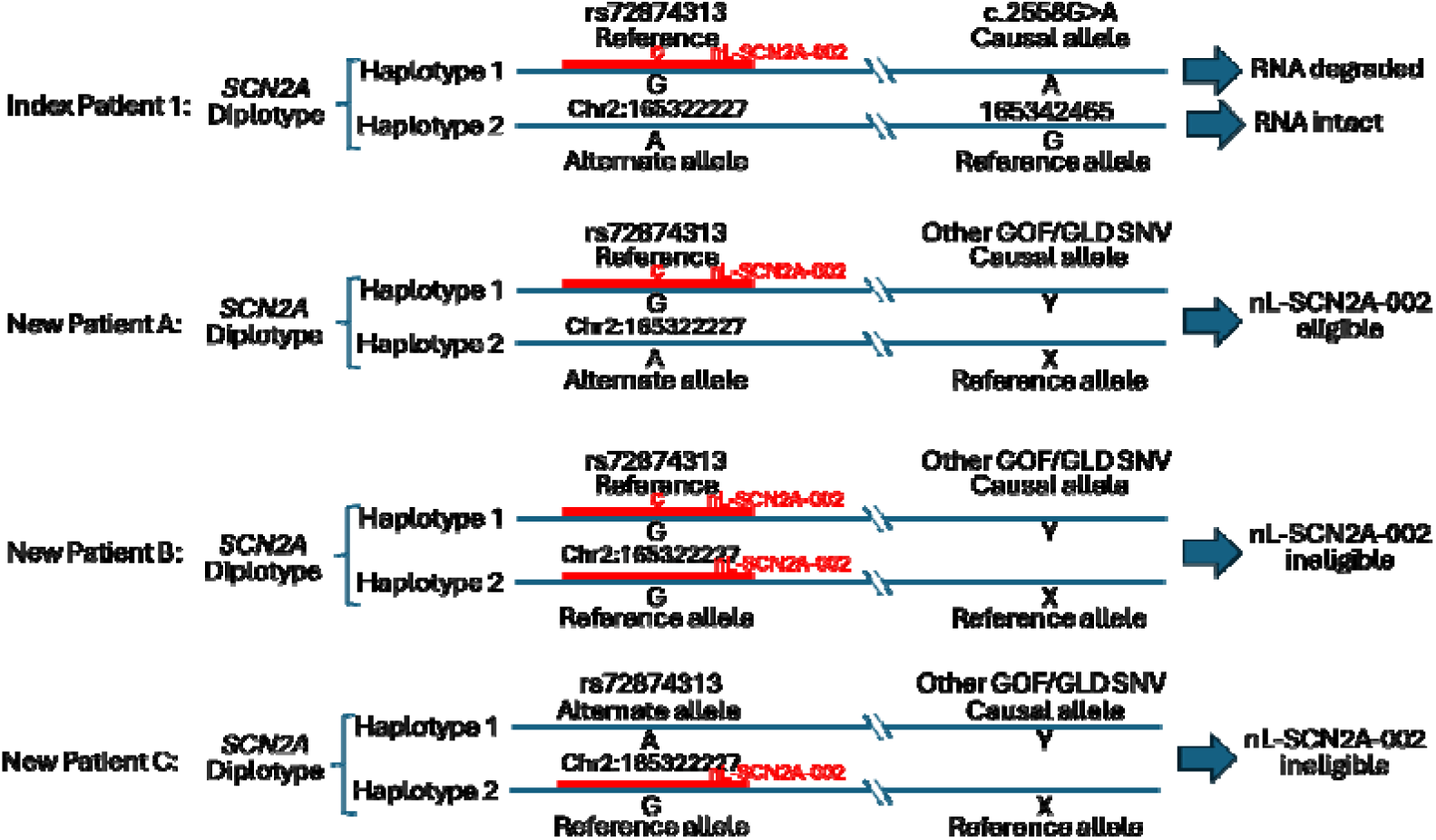
Diplotype-based expanded use of nL-SCN2A-002. *SCN2A* diplotypes (paired haplotypes, blue lines) are shown for four potential *SCN2A*-CND patients. In Index Patient 1, *SCN2A*-CND was caused by heterozygous c.2558G>A. nL-SCN2A-002 (red line) targeted 20 nt of reference *SCN2A* sequence (NC_000002.12:g.165322220-165322241=), 20,224 nt proximal to the causal variant. Selectivity for transcripts containing c.2558G>A was due to the patient being heterozygous for the common SNP rs72874313 (g.165322227A>G), located within the nL-SCN2A-002 target region, with the reference SNP allele *in cis* with the causal variant. nL-SCN2A-002 bound to and degraded *SCN2A* pre-mRNA with the haplotype g.[165322227=;165342465G>A] (rs72874313 reference; *SCN2A* c.2558G>A) preferentially to those with the haplotype g.[165322227A>G;165342465=] (rs72874313 alternate allele; *SCN2A* coding domain reference). New patient A with SCN2A-CND due to the same or a different gain-of-function (GOF) or mixed gain/loss dysfunction variant (*SCN2A* c.X>Y), would be potentially eligible for nL-SCN2A-002 if he or she had a diplotype with the causal *SCN2A* allele *in trans* to the heterozygous rs72874313 alternate allele. New patients B and C with SCN2A-CND due to the same or other different GOF or mixed gain/loss dysfunction variants, would not be eligible for nL-SCN2A-002 since they lacked haplotypes that would confer ASO specificity for the causal pre-mRNA.

At 30 mg intrathecal dosing, he experienced >90% seizure reduction and improved communication, irritability, ataxia, autonomic function, and walking. These were sustained over 1.5 years of treatment (Kim-McManus et al, submitted).

### Potential for expanded use of an individualized ASO for SCN2A-CND

Heterozygosity for rs72874313 is common (gnomAD 4.1 alternate allele frequency (altAF) 0.22). Thus, while developed for one child, patients with other *SCN2A*-CND causing variants could potentially benefit from nL-SCN2A-002 if they had compatible mechanisms and diplotypes (Fig. S1). Since most *SCN2A*-CND variants occur *de novo*, they are randomly associated with chromosome 2 haplotypes. Thus, ∼11% of individuals with *SCN2A*-CND were anticipated to be heterozygous reference for rs72874313 *in cis* with their causal variant, conferring nL-SCN2A-002 compatibility.

We evaluated phased WGS data from the 3,202 individuals of the 1000 Genomes Project (1kGP) to characterize the common haplotypes containing rs72874313.^33^ Using PLINK, rs72874313 mapped within haplotype block 1 of the *SCN2A* coding domain (NC-000002.12: g.165295823 - 165389822) (Fig. 1, S2). *SCN2A* haplotype block 1 was 30 kb in length (NC-000002.12: g.165296685-165326704) and was a region that contained many previously described *SCN2A*-CND causal variants. There were 109 distinct block 1 haplotypes in the 1kGP subjects, of which eight were present in >100 1kGP individuals (Table S1). In total, 914 1kGP individuals were heterozygous for rs72874313 across 21 haplotypes, with the majority (837 individuals) carrying the same common haplotype block: Hap1.3 (Fig. 1, Table S1). Thus, 28% of 1kGP subjects had potentially nL-SCN2A-002 compatible haplotypes.

**Fig. 1.**
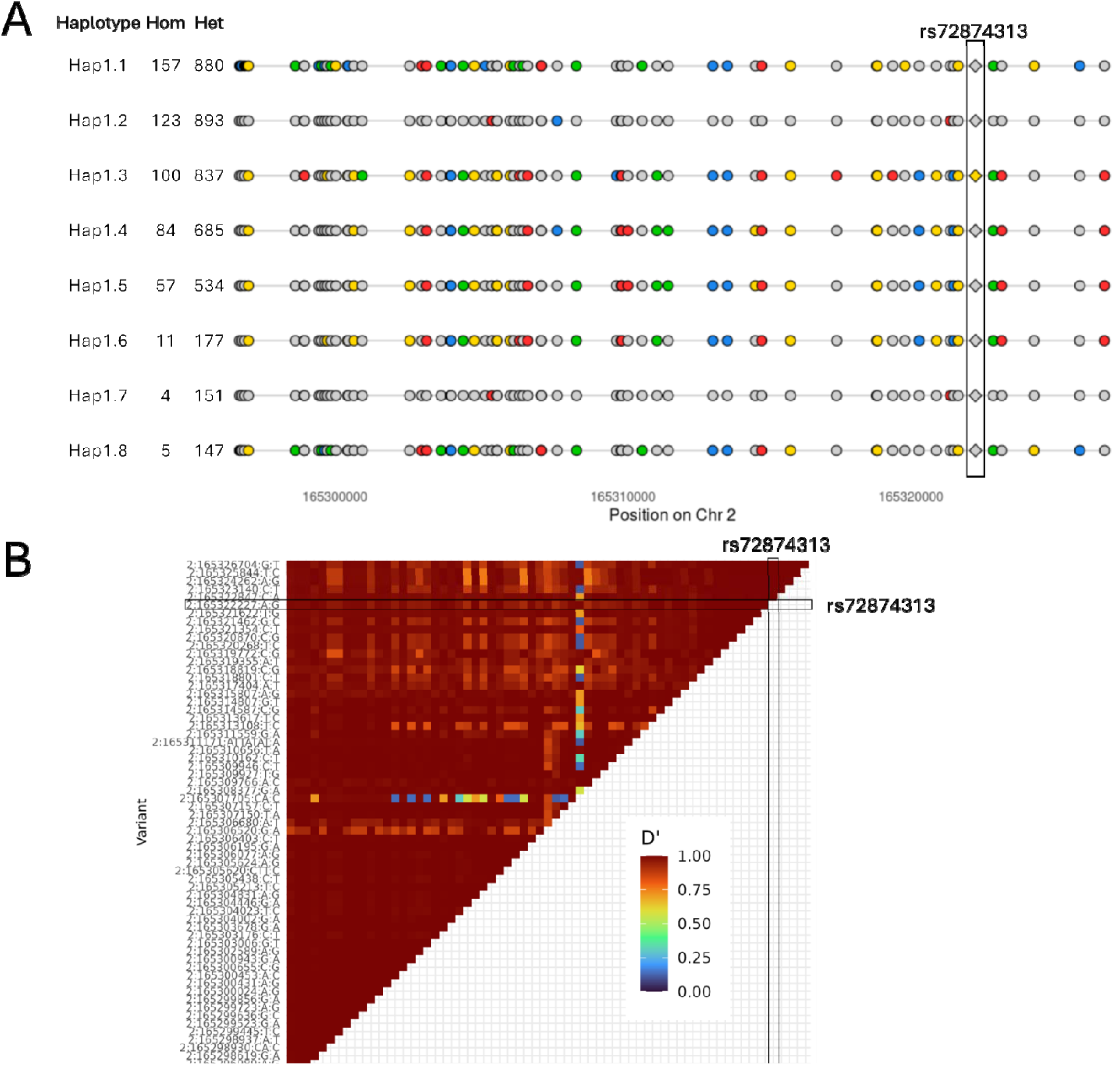
*SCN2A* haplotype block 1 determined by PLINK, showing haplotypes observed in ≥100 samples from the 1000 Genomes Project. (**A**) Haplotypes 1.1 to 1.8, showing numbers of homozygous (hom) and heterozygous (het) 1kGP individuals and positions and alleles of 66 informative single nucleotide polymorphisms (SNPs). Details are shown in Table S1. SNPs in light grey indicate reference alleles in the given haplotype, colors indicate the alternate allele: green – A, red – T, yellow – G, blue – C, black – indel. The SNP conferring allele specificity (rs72874313) to nL-SCN2A-002 is indicated by a diamond and is boxed. (**B**) Square linkage disequilibrium matrix (D’) for informative SNPs in th haplotype block.

**Figure S2:**
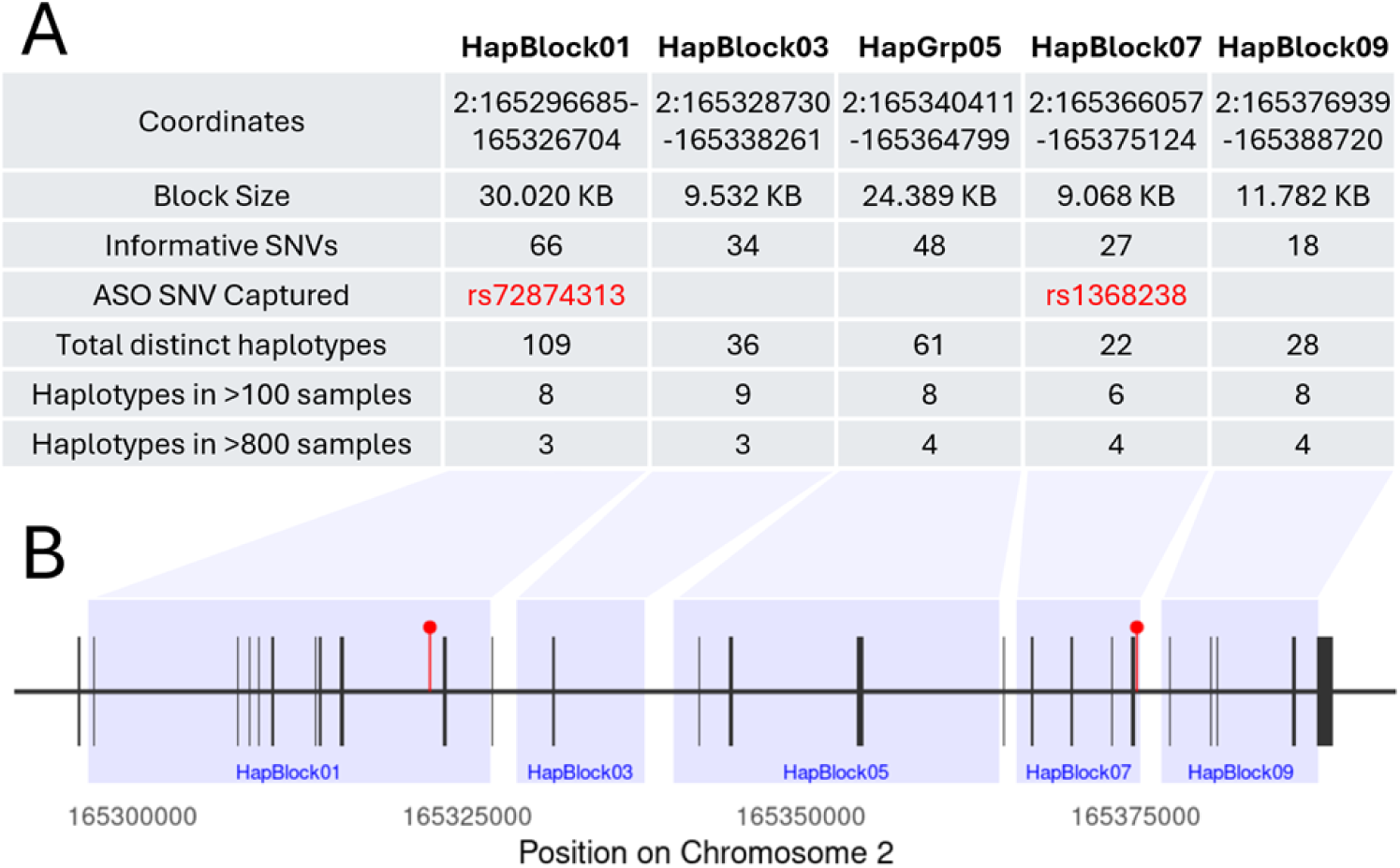
Overview of *SCN2A* haplotype blocks larger than 5 kb. (**A**) Table summarizing haplotype block results as detected by PLINK in the 1000 Genomes Project phased WGS: Chromosome (Chr) 2 GRCh38 coordinates, block size, informative single nucleotide polymorphisms (SNPs, altAF >0.15), total number of distinct haplotypes, and the subset of haplotypes observed in at least 100 distinct individuals. Only haplotype blocks >5 kb are included. Details are shown in Table S1 and S2. (**B**) Gene model of the *SCN2A* coding domain. Dark blocks indicate coding exons; red lollipops indicate ASO target SNPs; blue rectangles indicate the genomic boundaries of the haplotype blocks.

Given the frequency of haplotype-based eligibility for nL-SCN2A-002 in a healthy population, we evaluated nL-SCN2A-002 compatibility in twenty additional *SCN2A*-CND probands that we diagnosed by rapid clinical WGS from 2018–2024 (Table 1).^34^ Each had an unique causal variant. Effects of causal variants on neuronal Na+ channels were unknown in fourteen patients, three had Gain-of-Function (GOF) effects and two had mixed gain/loss dysfunction.^35–37^ The latter five were potential candidates since nL-SCN2A-002 had shown efficacy in a mixed gain/loss patient and ASOs are effective for GOF variants.

**Table 1:**
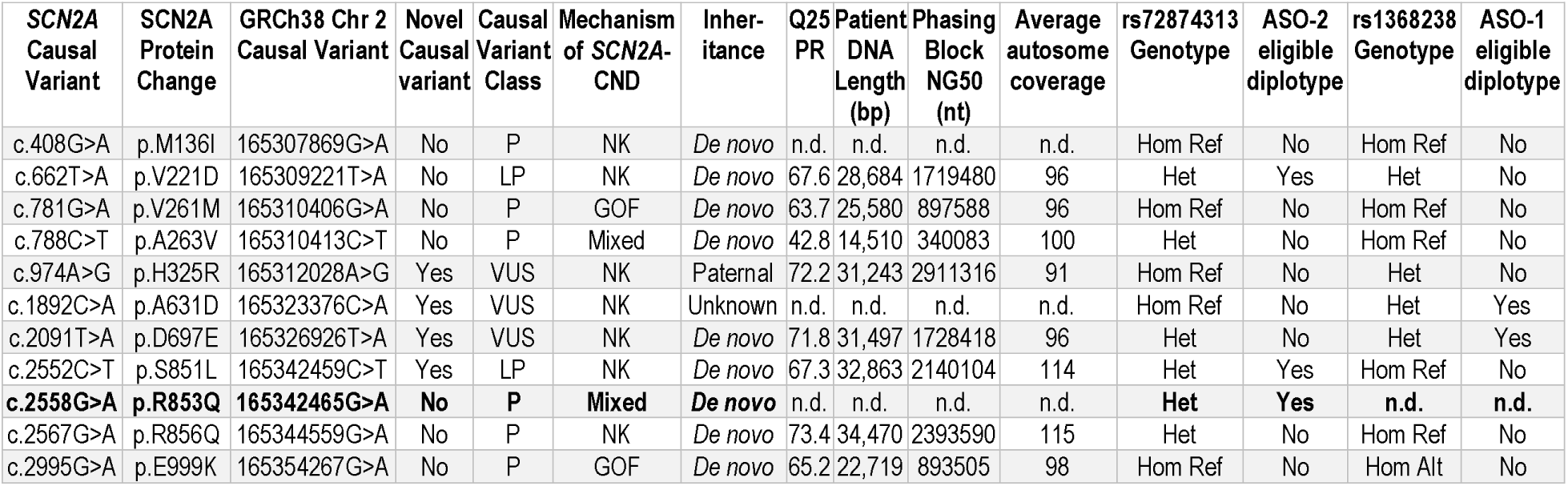
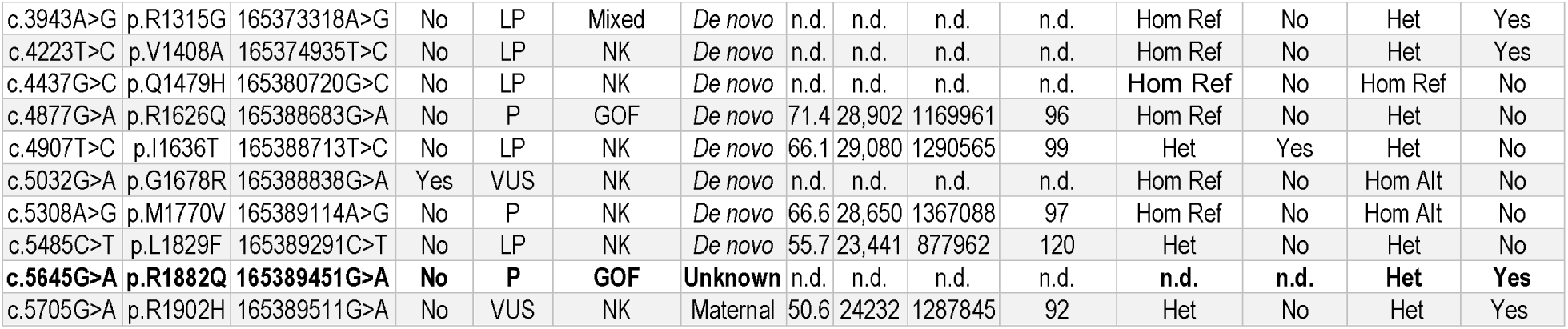
Diplotype-based evaluation of 21 *SCN2A*-CND patients for eligibility for ASOs nL-SCN2A-002 (ASO-2) and -001 (ASO-1) . Patients for whom the ASOs were developed are in bold. Abbreviations: Q25 PR, % read-pairs with Phred-like likelihood score >25 for genomically adjacent read-pair in neighboring nanowell; ACMG, American College of Medical Genetics; MOI, mode of inheritance; bp, base pairs; nt, nucleotides; NK, not known; n.d., not done.

WGS re-analysis revealed rs72874313 heterozygosity in eight of 21 *SCN2A*-CND probands (Table 1). The distance from rs72874313 to the causal variants ranged from 1.1 – 67 kb. Short-read WGS generates haplotypes of only a few kilobases, which is extensible to ∼20 kb by tagging SNPs in trios. However, the accuracy of haplotypes is poor since they are informed by only a few reads at bottleneck regions. Instead, we haplotyped the *SCN2A* genomic region using novel, short-read WGS with haplotyping by read proximity (Constellation research technology, Illumina).^38^ Unfragmented DNA was directly loaded onto the flow cell where surface-bound transposomes bound and performed tagmentation *in situ*, followed by standard Illumina clustering and sequencing (Fig. 2a). Thus, adjacent nanowells were likely to be occupied by fragments derived from the same DNA molecule. Assembly utilized the physical location of reads on the flow cell to create spatially proximal sets of reads derived from contiguous genomic DNA molecules (DRAGEN, Illumina) and enabled the reconstruction of multi-megabase haplotypes.^39^ Twenty-one *SCN2A*-CND patients underwent Constellation WGS. Each lane of a NovaSeq X Plus 10B flowcell was loaded with 350 ng of genomic DNA of an *SCN2A*-CND patient (average length 23.4 – 34.5 kb, Table 1). Paired 151-cycle WGS achieved median autosomal coverage of 97-fold.

**Fig. 2.**
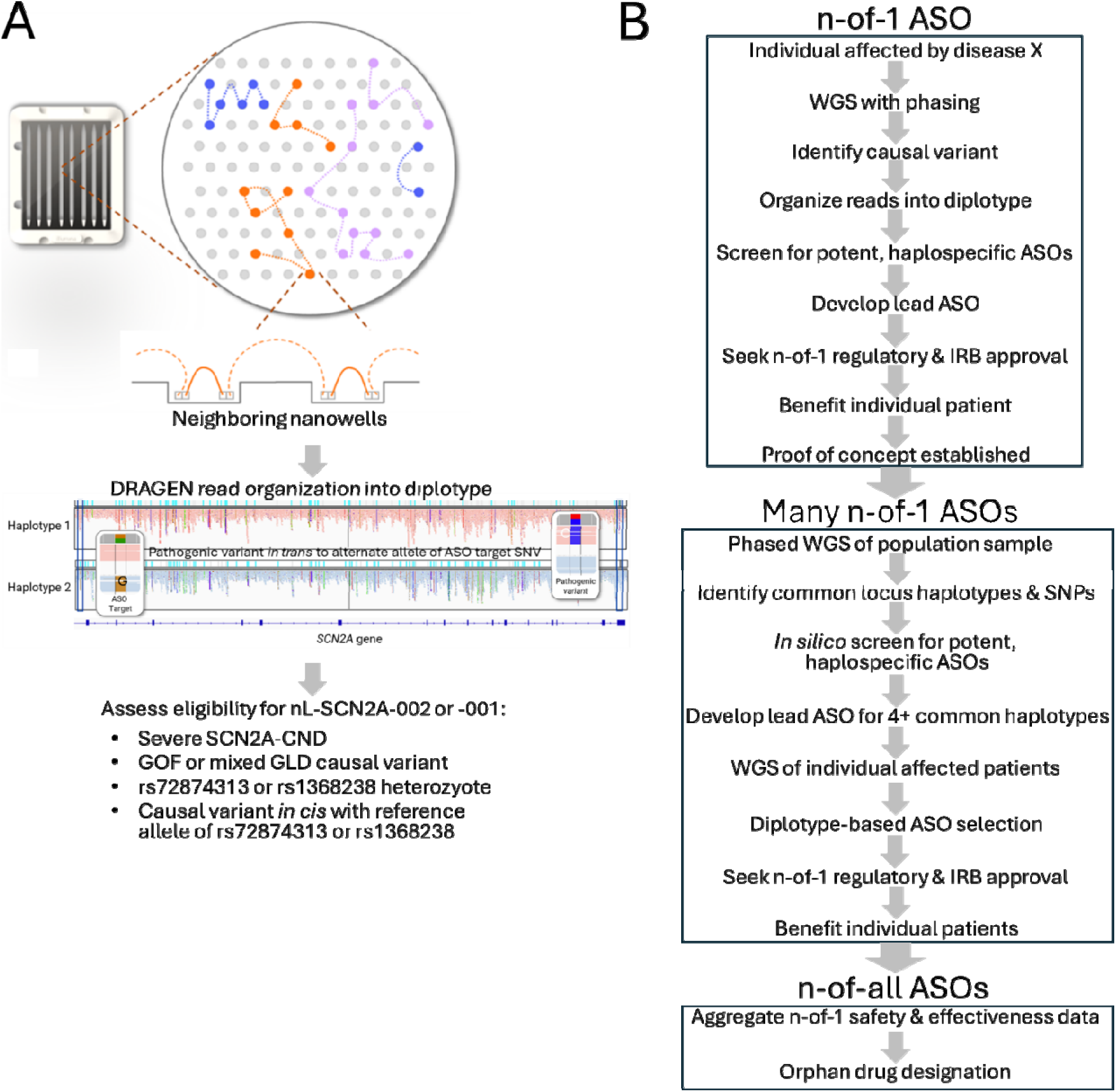
A diplotype-based method for expanded use of haplospecific ASOs. (**A**) Constellation WGS provides *SCN2A* phasing to assess diplotype compatibility of *SCN2A*-CND patients for nL-SCN2A-002 or -001. (**B**) An efficient pathway for development of a set of haplospecific ASOs for most patients with a genetic disorder. Following development and successful use of a first haplospecific ASO in an index patient, population haplotyping is undertaken to identify all common haplotypes, a set of haplospecific ASOs, and their evaluation in several n-of-1 trials. Safety and effectiveness data from several n-of-1 trial is aggregated and subsequently used for orphan drug-designated ASOs for each common haplotype.

Two thirds of read-pairs had an adjacent nanowell with a genomically adjacent read-pair (median Phred-scaled quality score >25 proximity rate 66.6%, range 42.8 – 73.4%). This generated a median phased block NG50 of 2.1 Mb (range 0.9 – 2.4 Mb). The proximity rate and NG50 varied linearly with the average input genomic DNA length. The *SCN2A* locus was fully phased in all samples (Fig. 2a). Four patients had nL-SCN2A-002–compatible diplotypes (Table 1, Fig. S1). While none of the associated causal variants in these patients (SCN2A p.S851L, p.V221D, and p.I1636T) had been characterized functionally, all induced neonatal-onset seizures which mechanistically are strongly associated with *SCN2A* GOF variants.^35^

### N-of-1 use of a second ASO for SCN2A-CND

The generalizability of a diplotype-based n-of-1 to n-of-many therapeutic pathway was examined in a second ASO (nL-SCN2A-001, n-Lorem). It was also a 20 nt phosphorothioate 2’methoxyethyl gapmer. It targeted reference sequence of *SCN2A* intron 23 (NC_000002.12: g.165375116-165375135=). It showed safety and efficacy in an individualized trial in a boy with *SCN2A*-CND and medically refractory seizures of neonatal onset, ASD and severe ID (Kim-McManus et al, submitted). His causal variant (*SCN2A* c.5645G>A, p.R1882Q) had GOF effect. The causal variant was located 14 kb from the ASO target sequence. nL-SCN2A-001 selectivity was due to his heterozygosity for rs190030016 (g.165375126A>G), which lay within the nL-SCN2A-001 target sequence, and his causal *SCN2A* haplotype of g.[165375126=; 165389451G>A] (rs190030016 reference allele; SCN2A p.R1882Q) (Fig. 1a, S3). Intrathecal bimonthly 40–60 mg dosing led to sustained reduction in seizures and abnormal behavior, and improvement in neurodevelopmental skills for 2 years.

**Figure S3:**
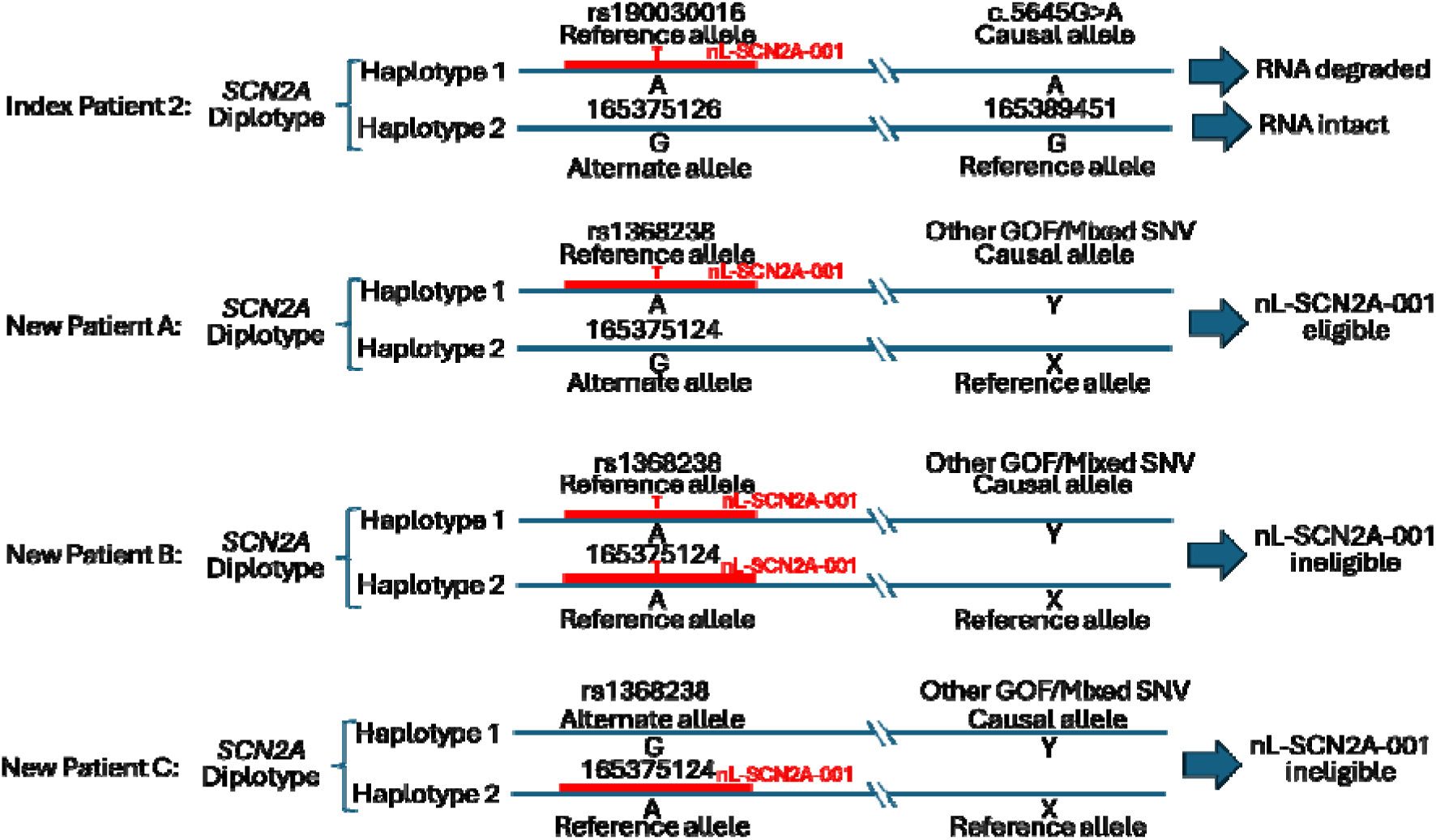
Diplotype-based expanded use of nL-SCN2A-001. *SNC2A* diplotypes (paired haplotypes, blue lines) are shown for four *SCN2A*-CND patients. In Index Patient 2, *SCN2A*-CND was caused by heterozygous c.5645G>A (NC_000002.12:g.165389451G>A). nL-SCN2A-001 (red line) targeted reference *SCN2A* sequence (NC_000002.12:g.165375116-165375135=), 14,316 nt proximal to the causal variant. Selectivity for transcripts containing c.5645G>A was due to the patient being heterozygous for the rare SNV rs190030016 (g. 165375126A>G, which is located within the nL-SCN2A-001 target region) with the reference allele *in cis* with the nL-SCN2A-001 target sequence. nL-SCN2A-001 bound to and degraded *SCN2A* pre-mRNA with the haplotype g.[165375126=;165389451G>A] (rs190030016 reference and *SCN2A* c.5645G>A) preferentially to those with the haplotype g.[165375126A>G;165389451=] (rs190030016 alternate allele and *SCN2A* coding domain reference). A common SNP (rs1368238, g.165375124A>G) was also located in the nL-SCN2A-001 target sequence. New patient A with *SCN2A*-CND due to the same or a different gain-of-function (GOF) or mixed gain/loss dysfunction variant (*SCN2A c.*X>Y), would be potentially eligible for nL-SCN2A-002 if he or she had a diplotype with the causal *SCN2A* allele *in trans* to the heterozygous rs1368238 alternate allele. New patients B and C with *SCN2A*-CND due to the same or other different GOF or mixed gain/loss dysfunction variants (*SCN2A*X>Y), would not be eligible for nL-SCN2A-001.

### Potential for expanded use of a second individualized ASO for SCN2A-CND

rs190030016 was too rare (altAF 0.7/100,000) for other *SCN2A*-CND patients to be eligible. However, a common SNP (rs1368238, g.165375124A>G, altAF 0.29) was also located in the nL-SCN2A-001 target sequence (Fig. S3). Thus, *SCN2A*-CND patients with diplotypes containing causal variants *in cis* with the heterozygous reference allele of rs1368238 could potentially benefit from nL-SCN2A-001 (Fig. 1a, S3).

In the 3,202 1kGP subjects, rs1368238 mapped within haplotype block 7 of the *SCN2A* coding domain. *SCN2A* haplotype block 7 was 9.1 kb in length (NC-000002.12: g.165366057-165375124) but also contained many *SCN2A*-CND causal variants (Fig. 3). There were 22 distinct block 7 haplotypes in the 1kGP subjects, of which 6 were present in >100 1kGP individuals (Fig. 3, Fig. S2, Table S1). Four haplotypes contained the alternate allele for rs1368238, contributing to 1,230 out of 3,202 1kGP subjects heterozygous for rs1368238; one haplotype 7 block, Hap7.1, accounted for 1,216 of these heterozygous individuals (Table S1). Thus, 38% of 1kGP subjects had potentially nL-SCN2A-001 compatible haplotypes, of which one half (19%) would be expected to have a causal variant *in cis*.

**Fig 3.**
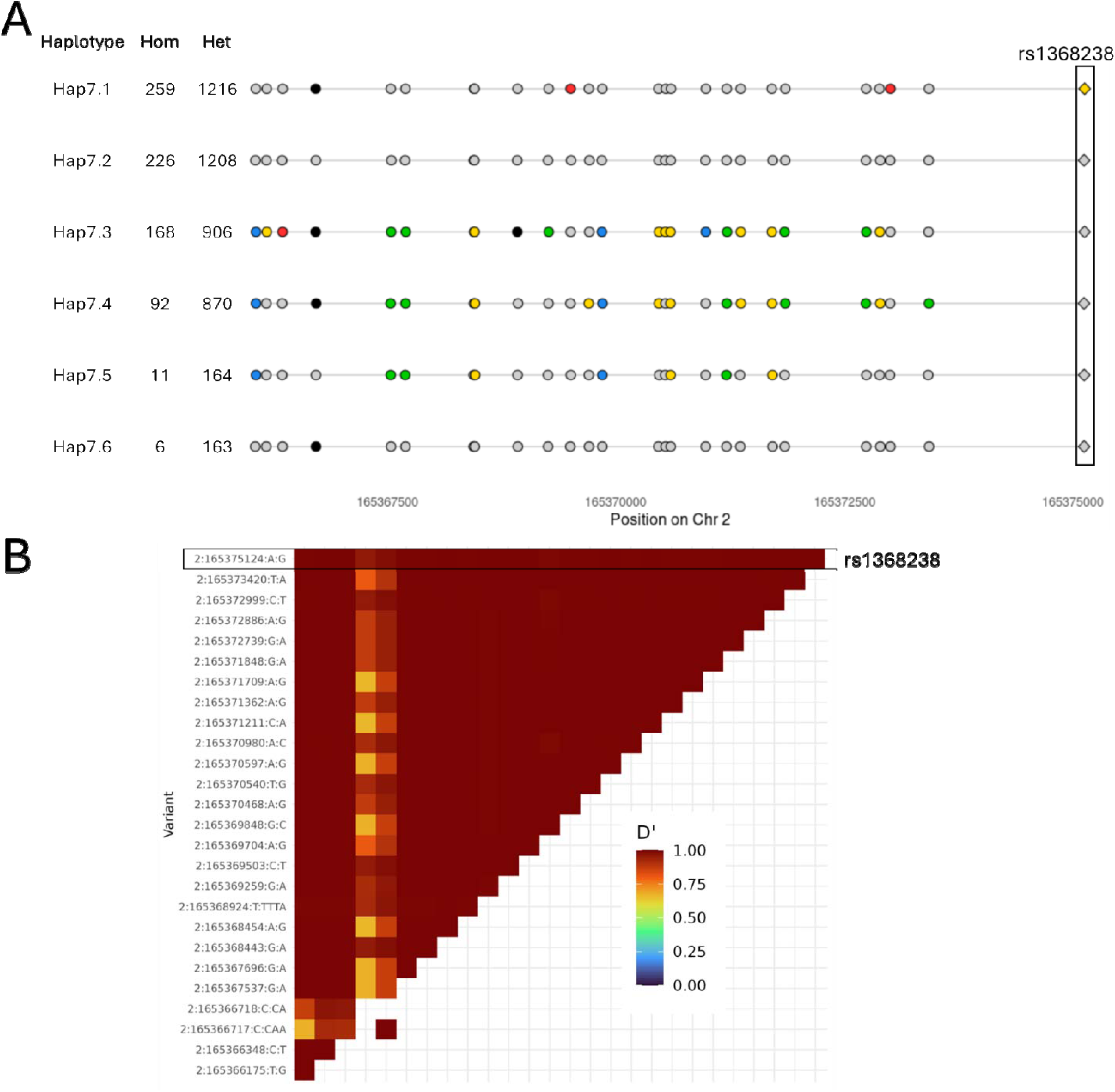
*SCN2A* haplotype block 7 determined by PLINK, showing haplotypes observed in ≥100 samples from the 1000 Genomes Project. (**A**) Haplotypes 7.1 to 7.6, showing numbers of homozygous (hom) and heterozygous (het) 1kGP individuals and positions and alleles of 26 informative single nucleotide polymorphisms (SNPs). SNPs in light grey indicate reference alleles in the given haplotype, colors indicate the alternate allele: green – A, red – T, yellow – G, blue – C, black – indel. The SNP conferring allele specificity (rs1368238) to nL-SCN2A-001 is indicated by a diamond and is boxed. (**B**) Square linkage disequilibrium matrix (D’) for informative SNPs in the haplotype block. The SNP conferring allele specificity (rs1368238) to nL-SCN2A-001 is boxed.

Constellation WGS revealed that six of 21 *SCN2A*-CND patients could potentially benefit from nL-SCN2A-001 (Table 1). Of the five additional *SCN2A*-CND variants, only one has been characterized functionally: p.R1315G is characterized by mixed gain/loss dysfunction. This patient had neonatal-onset seizures at time of diagnosis by WGS. *In toto*, nL-SCN2A-002 and - 001 had potential applicability in ten of 21 *SCN2A*-CND patients (Table 1).

### Haplotype-informed selection of additional individualized ASOs for expanded use

For *SCN2A*-CND patients with ineligible diplotypes for nL-SCN2A-002 and -001 we sought additional candidate *SCN2A* allele-specific ASOs that could be suitable for N-of-1 trials in any future patient with a pathogenic variant conferring GOF or mixed dysfunction. We identified common SNPs that tagged haplotype blocks other than Hap001.3 and Hap007.4, and that could confer allele specificity when located within an ASO target sequence. The 94 kb *SCN2A* coding domain contained nine haplotype blocks in the 1kGP subjects, of which five were >5 kb in length and contained 190 common SNPs (altAF >0.15) (Fig. 1, 3, S2, S4 – S6, Table S2).

**Fig. S4.**
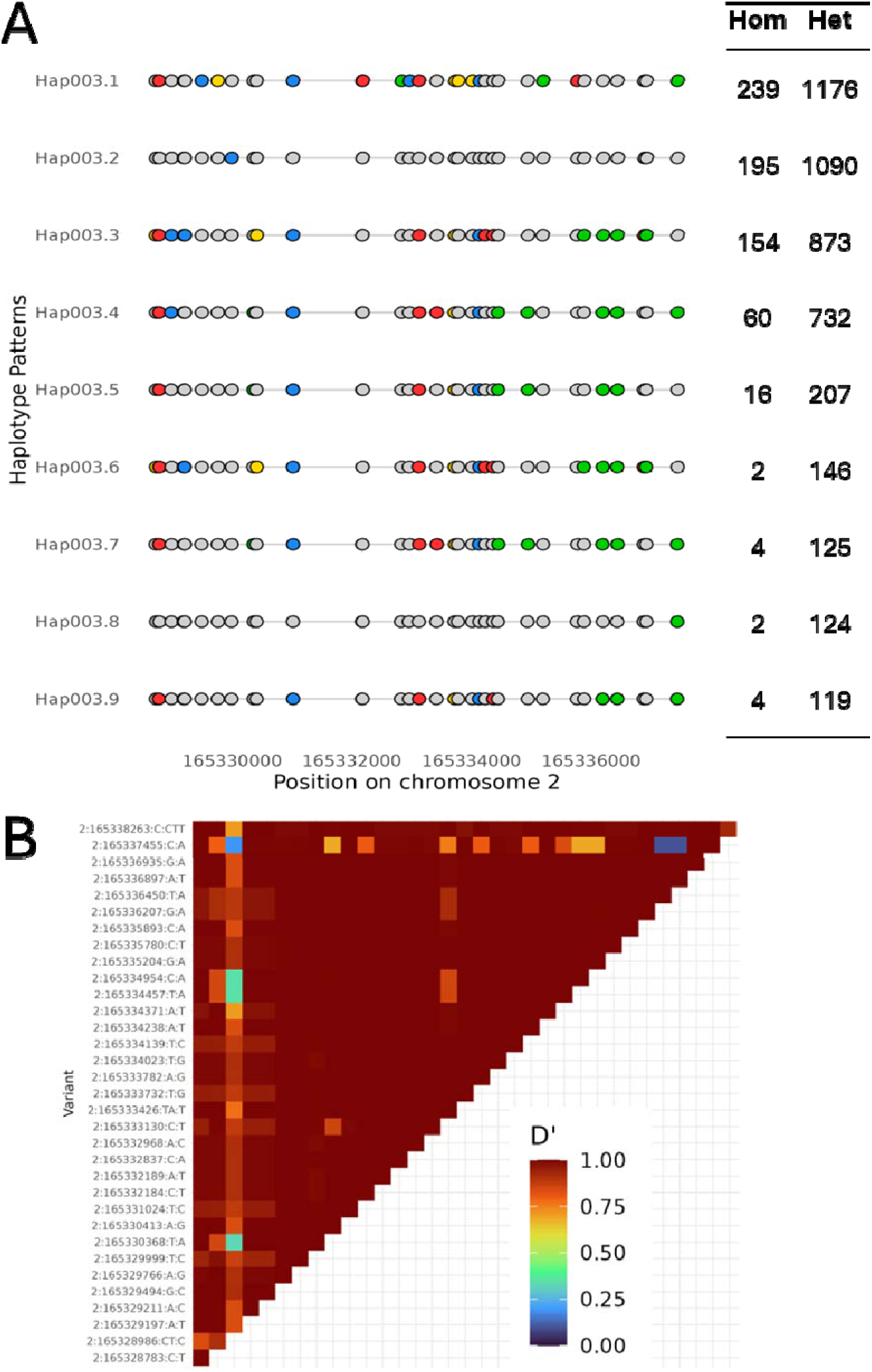
*SCN2A* haplotype block 3 determined by PLINK, showing haplotypes observed in ≥100 samples from the 1000 Genomes Project. (**A**) Haplotypes 3.1 to 3.9, showing numbers of homozygous (hom) and heterozygous (het) 1kGP individuals and positions and alleles of 34 informative SNPs. SNPs in light grey indicate reference alleles in the given haplotype, colors indicate the alternate allele: green – A, red – T, yellow – G, blue – C, black – indel. (**B**) Square linkage disequilibrium matrix (D’) for informative SNPs in the haplotype block.

**Fig. S5.**
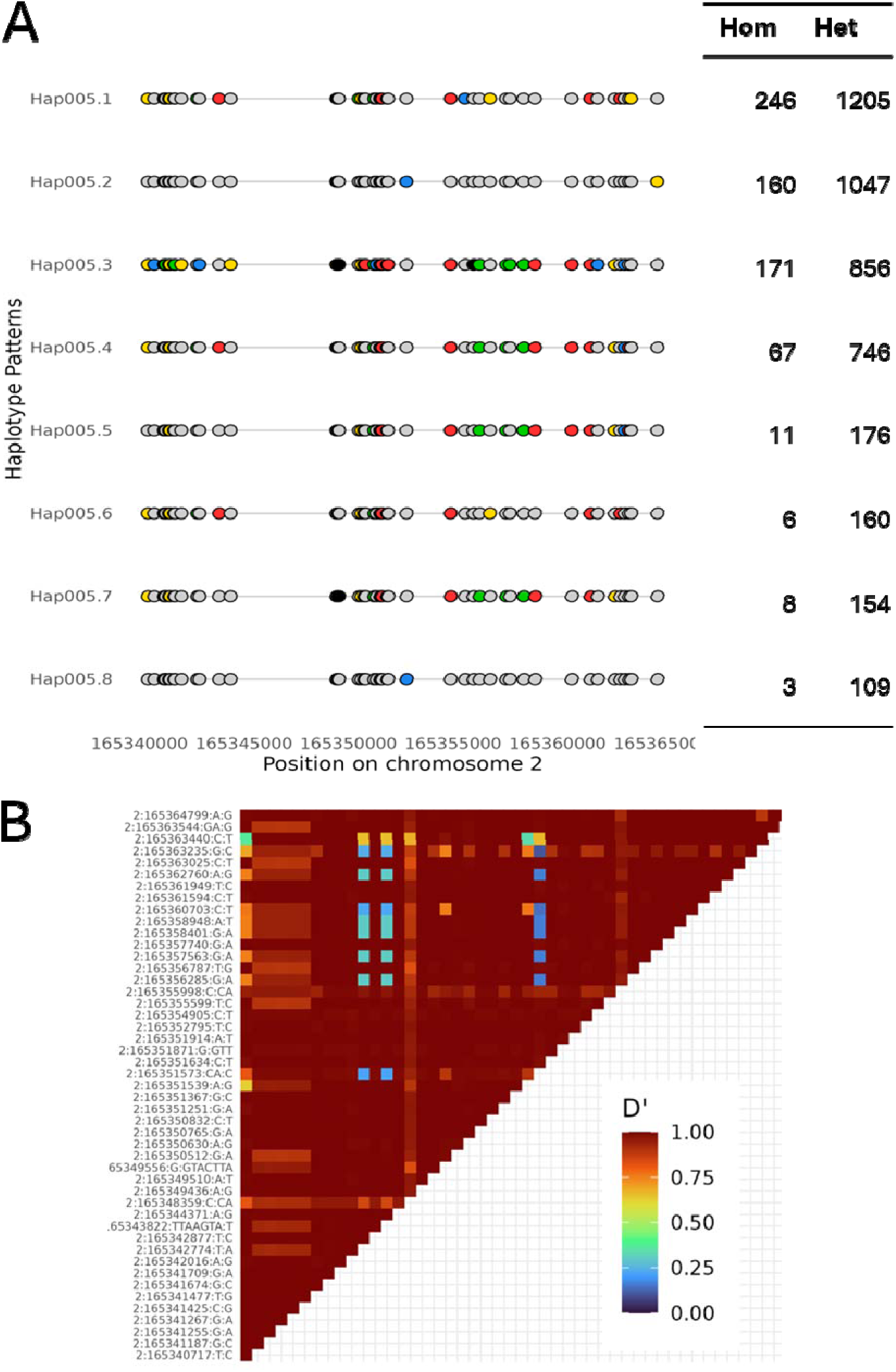
*SCN2A* haplotype block 5 determined by PLINK, showing haplotypes observed in ≥100 samples from the 1000 Genomes Project. (**A**) Haplotypes 5.1 to 5.8, showing numbers of homozygous (hom) and heterozygous (het) 1kGP individuals and positions and alleles of 48 informative SNPs. SNPs in light grey indicate reference alleles in the given haplotype, colors indicate the alternate allele: green – A, red – T, yellow – G, blue – C, black – indel. (**B**) Square linkage disequilibrium matrix (D’) for informative SNPs in the haplotype block.

**Fig. S6.**
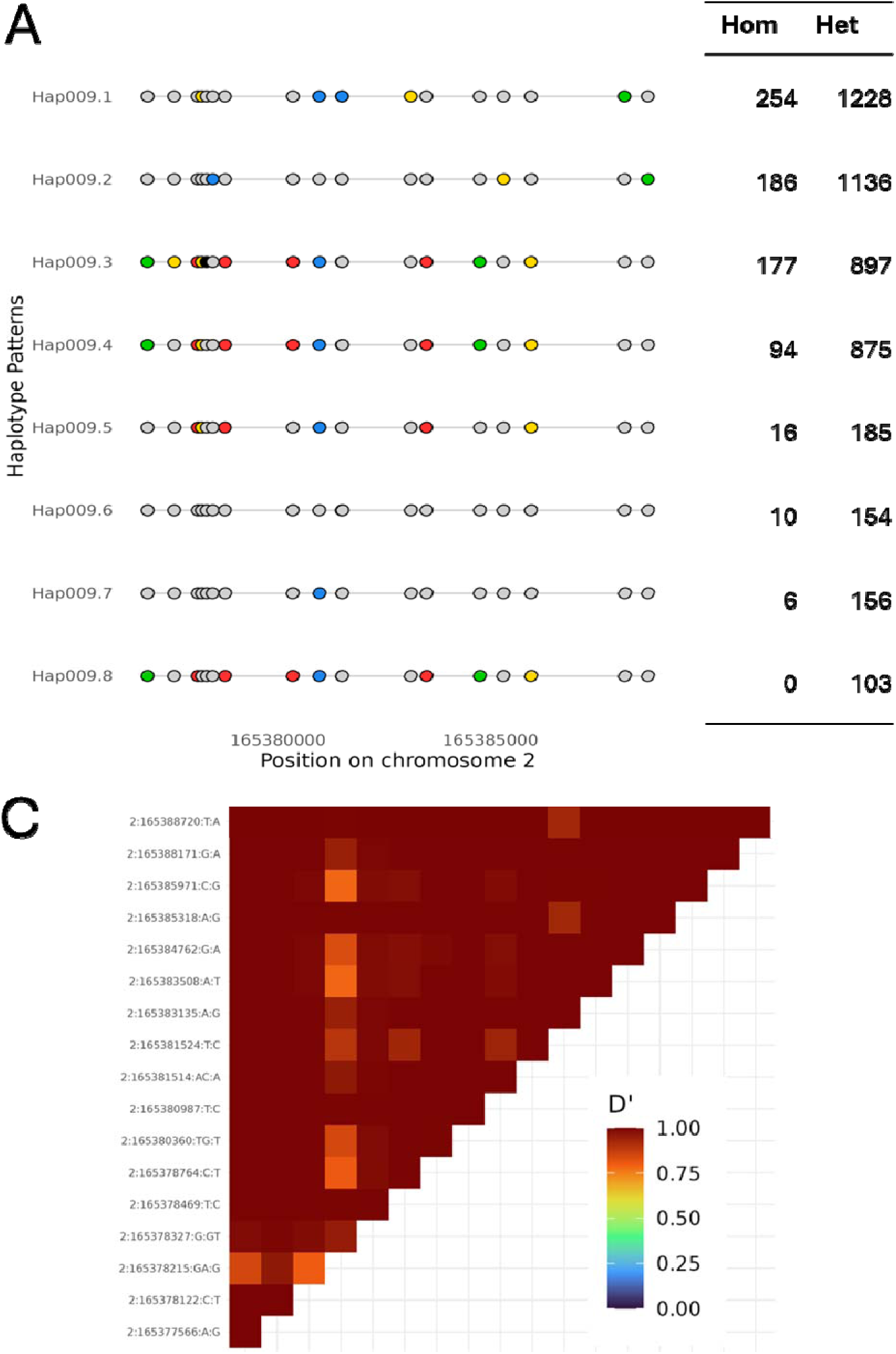
*SCN2A* haplotype block 9 determined by PLINK, showing haplotypes observed in ≥100 samples from the 1000 Genomes Project. (**A**) Haplotypes 9.1 to 9.8, showing numbers of homozygous (hom) and heterozygous (het) 1kGP individuals and positions and alleles of 18 informative SNPs. SNPs in light grey indicate reference alleles in the given haplotype, colors indicate the alternate allele: green – A, red – T, yellow – G, blue – C, black – indel. (**B**) Square linkage disequilibrium matrix (D’) for informative SNPs in the haplotype block.

While the five haplotype groups were represented by 256 distinct haplotypes in the 1kGP subjects, only 18 haplotypes were present in >800 (25%) 1kGP subjects (Fig. S2). In theory, broad coverage of *SCN2A*-CND causal variants could efficiently be achieved with a small set of ASOs defined by selectivity for the most common haplotypes. After excluding the 37 common SNPs whose alternate alleles were associated with haplotype blocks Hap001.3 and Hap007.4, 156 SNPs representing 16 haplotypes were targets for additional allele-specific *SCN2A* ASOs (Table S1, S2). Additional candidate haplospecific *SCN2A* ASOs containing these SNPs and with physicochemical properties conferring high likelihood of *in vivo* potency and specificity were selected by *in silico* modelling of reference Chr 2 sequences (Table 2, S3)^40–44^. Two were modifications of nL-SCN2A-001 and -002 with complementarity to the alternate alleles of rs1368238 and rs72874313, respectively (RC-001 and RC-002, Table S3). A combination of four ASOs was sufficient for eligibility for 2,444 (76%) 1kGP subjects (Table 2). Of 21 *SCN2A*-CND patients, 15 were eligible for one of four ASOs (nL-SCN2A-001 or -002 or RC-001 or -002 (Table 1).

**Table 2:**
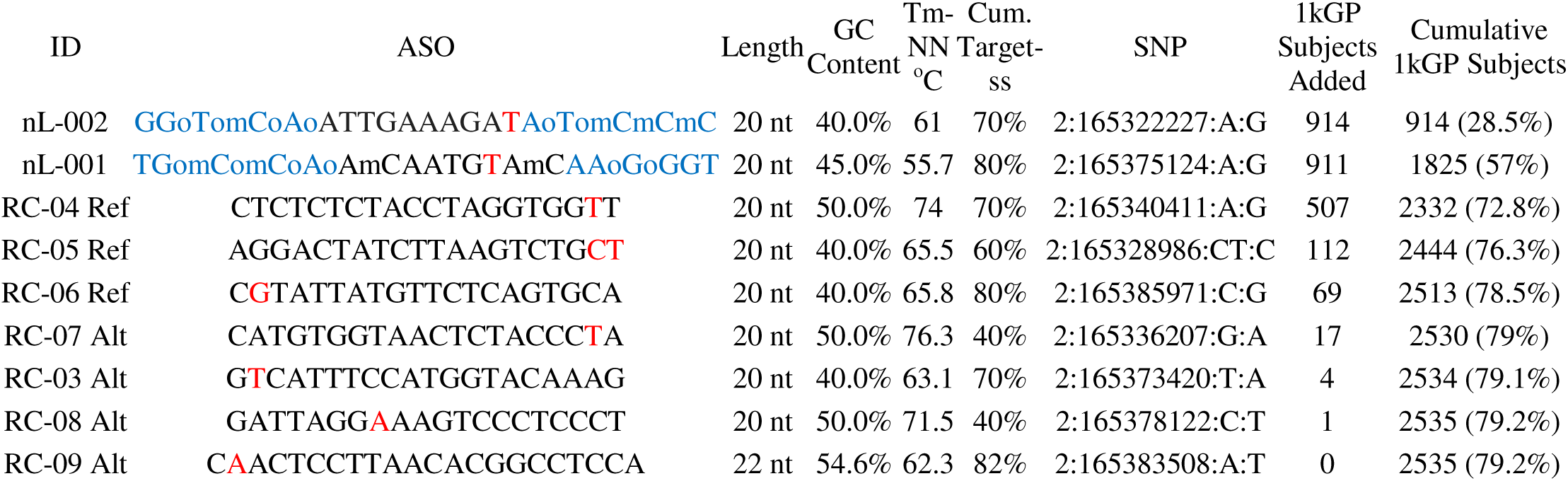
Cumulative eligibility of 1kGP subjects by a panel of *SCN2A* haplospecific ASO candidate sequences. . For nL-001 and nL-002 blue nucleotides are 2’ methoxyethyl deoxyribose, unmarked backbone linkages were phosphorothioate, linkages marked with o were normal phosphodiester, and mC was 5-methylcytosine. Tm-NN, melting temperature using a nearest-neighbor thermodynamic model; Cum. Target-ss, cumulative percentage of target-sense RNA strands being successfully modulated (silenced/modified) by an ASO.

1kGP lymphoblastoid cell lines with heterozygosity for the tagging SNPs were identified (Table S2). Following reprogramming to induced pluripotent stem cells and differentiation into *SCN2A*-expressing neural progenitors these could provide measurements for *in vitro* potency and allele selectivity of novel ASOs^45,46^. In this manner, a set of haplospecific ASOs with predetermined *in vitro* potency and allele selectivity would be pre-made for most future *SCN2A*-CND patients, with ASO selection by *SCN2A* haplotyping at time of diagnosis.

## DISCUSSION

Here we described a pathway to extend the utility of two individualized ASOs from initial N-of-1 use in index patients to their readiness for evaluation in N-of-1 trials in approximately one half of children affected by *SCN2A*-CND, a severe childhood-onset disorder that currently lacks disease modifying therapy. We also described methods that used 1kGP resources to pre-design a set of four haplospecific ASOs with potential utility in 76% of children with *SCN2A*-CND.

*SCN2A*-CND is an ideal disease candidate for this N-of-1 to broad use pathway for several reasons. Firstly, it is a relatively common cause of epileptic encephalopathy and severe autism spectrum disorder. Secondly, it is effectively identified by WGS either at birth via WGS-based newborn screening or diagnostic WGS following symptom onset. Thirdly, the mechanism of disease is well understood for several *SCN2A* disease phenotypes, and *in vitro* assays exist to determine the mechanism for novel variants. Fourthly, the rate of progression of *SCN2A*-CND is sufficiently slow to allow time to prepare N-of-1 regulatory packets. Fifthly, demonstration of safety and effectiveness of two ASOs in index patients greatly improves the benefit:risk likelihood in future patients with *SCN2A* diplotypes compatible for use with nL-SCN2A-001 and -002. In aggregate they had broad potential for *SCN2A*-CND treatment since they have shown efficacy for several phenotypes, two functional mechanisms, and target common haplotypes. In addition, experience in two patients has validated a delivery route, effective therapeutic ranges, frequency of administration, and sets of outcome and safety measures. Likewise, their safety and effectiveness improve the benefit:risk ratio for novel haplospecific ASOs in *SCN2A*-CND. A possible pathway from N-of-1 to broad applicability would be to seek FDA authorization for nL-SCN2A-002 and -001 use in additional patients with compatible diplotypes and GOF or mixed gain-loss dysfunction variants. Upon demonstration of safety and efficacy in those individuals, the process would be reiterated to generate substantive in-human evidence to support orphan drug designation for these ASOs.

This approach is not limited to *SCN2A* or GOF and mixed dysfunction mechanisms. Consensus guidelines have been developed for assessing the eligibility of pathogenic DNA variants for ASO treatments^47^. For example, the individualized ASO nl-KIF1-001 was developed for one patient with *KIF1A*-associated neurological disorder (KAND) due to dominant negative heterozygous c.914C>T.^48^ However, nl-KIF1-001 targets reference intronic *KIF1A* sequences that harbor the common variant rs7578279 (altAF 0.54). Other patients with KAND could potentially benefit from nl-KIF1-001 if they have diplotypes comprising dominant negative causal variants *in cis* with the heterozygous rs7578279 reference allele.

An efficient, even more general approach for pre-development of haplospecific ASOs for rare, severe genetic disease populations is to ascertain *a priori* the complete set of common haplotypes for each disease locus in a well characterized population, such as the 1kGP (Fig. 1b). Herein we demonstrated a proof-of-concept for *a priori* haplotype-informed ASO design for *SCN2A*-CND. The 1kGP is large (3,202 individuals), with representation of human population diversity, and has both phased genome sequences and available lymphoblastoid cell lines for *in vitro* analyses. After identification of all common SNPs that tag each common haplotype, all haplospecific ASO candidates that target genomic regions encompassing these SNPs variants would be modeled *in silico* for stability, potency (optimal target sequence accessibility), and specificity (minimal off-target binding). Software tools exist that allow this to be undertaken as scale^40–44^. Resultant candidates would be synthesized and screened *in vitro* for potency and allele selectivity in 1kGP samples with appropriate SNP genotypes, and safety assessed in preclinical models. The efficiency of ASO pre-development could be further increased by routinely screening modifications of haplospecific ASOs that have already shown evidence in human of safety and effectiveness. Here we demonstrated this for nL-SCN2A-001 and -002 by screening for ASOs complementary to the alternate alleles of rs1368238 and rs72874313, respectively (RC-001 and RC-002). The latter differ chemically from the former by only one nucleotide and base and linkage modifications can be preserved. This would further increase the likelihood of safety in human. In addition, rank ordering the large number of informative SNPs for *SCN2A* would prioritize evaluation of those with the highest number of heterozygous 1kGP subjects, rather than the >800 (25%) cutoff used herein. For example, 48 SCN2A SNPs were heterozygous in >45% of 1kGP subjects.

These adaptations would be much more efficient and timely than the process used for nL-SCN2A-001 and -002. Absent knowledge of patient haplotypes, the latter each required synthesis of over 500 2’-MOE/DNA gapmer ASO candidates, and *in vitro* screening in patient-derived iPSC lines for potency and specificity. For nL-SCN2A-002 this delayed treatment by two years. As exemplified in the 1kGP set, it is possible to pre-qualify a small set of ASOs that tag different common haplotypes that together encompass most *SCN2A*-CND causal variants. Using 1kGP cell lines, these candidate ASOs could be evaluated *in vitro* before patient identification or derivation of a patient cell line. Upon diagnosis of affected individuals by WGS with concomitant haplotyping, such as by Constellation, the patient diplotype would allow selection of a suitable ASO candidate from the set (Fig. 1b). In this manner, much of the evidence for an FDA N-of-1 regulatory submission would be developed *a priori*. In the setting of N-of-1 aggregation, prior N-of-1 trials would provide additional evidence of safety and effectiveness for a genetic therapy platform, specific disorder, disease mechanism, delivery route, dose intensity, and set of outcome measures were aggregated to attempt to generate general evidence.^5,6^

Many severe childhood monogenic disorders result from pathogenic GOF variants and would potentially be amenable to pre-derivation of haplospecific ASO sets. In addition to *SCN2A*-CND, they include many cases of *SCN8A*-Developmental and epileptic encephalopathy (DEE) 13 and *KCNQ2*-CND, *KCNK3*-Developmental delay with sleep apnea, *NALCN*-Congenital contractures of the limbs and face, hypotonia, and developmental delay (CLIFAHDD) syndrome, *KCNJ11*-and *ABCC8*-Permanent neonatal diabetes mellitus, *PIK3CA*, *AKT1*, *MTOR*, and *CCND2* overgrowth syndromes, *HRAS*, *BRAF*, *MAP2K1*, *MAP2K2*, *KRAS*, and *NRAS* rasopathies, *TMEM173, SAVI, TREX1, IFIH1 JAK1, JAK2, STAT1, STAT3, NLRC4*, and *NLRP3* hyperinflammatory syndromes, *C3, CFB*, and *CFI* Atypical hemolytic uremic syndrome, and MPL and THPO myoproliferative states.^49–64^ Likewise, many severe childhood monogenic disorders result from pathogenic dominant negative variants and would also potentially be amenable to pre-derivation of haplospecific ASO sets. In addition to *KIF1A*-KAND, they include many cases of *SPTLC1*-Juvenile amyotrophic lateral sclerosis 27, ATP5F1A-Nuclear mitochondrial complex V deficiency 4A, *CARD11*-Immunodeficiency 11B with atopic dermatitis, *RELA*-Behcet-like familial autoinflammatory disease 3, *SPTAN1*-DEE5, and *KIF5B*-, *COL1A1*-, *COL1A2*-, and *COL2A1*-Osteogenesis imperfecta.^65–71^ Pre-derivation of haplospecific ASO sets for homogeneous clinical presentations would provide economies of scale. When combined with WGS for individuals with those presentations, diagnosis would allow selection of a haplospecific ASO for abbreviated N-of-1 development.

This N-of-1 to broad applicability approach would, in theory, be superior to use of a single non-selective ASO for treatment of a severe genetic disease for several reasons. Firstly, allele specific ASOs have superior safety by virtue of sparing normal transcripts from degradation. Secondly, by amenability for N-of-1 trial approval, time to treatment for individual patients is greatly accelerated. Thirdly, development risk is greatly diminished by this agile pathway relative to a waterfall approach to drug development.

In summary, individualized trials of diplotype-specific ASOs featuring common variants may be an efficient means to accelerate and de-risk drug development for severe, rare genetic diseases – immediately benefitting individual patients while generating human safety and efficacy evidence that informs subsequent orphan drug designation.

## MATERIALS AND METHODS

### Subjects and Ethics

All work was conducted with ethics/IRB approval by the Western Institutional Review Board / Copernicus Group (WCG Clinical) and performed in accordance with the Declaration of Helsinki. N-of-1 clinical trials were approved by the Institutional Review Board at the University of California-San Diego and the Food and Drug Administration (ClinicalTrials.gov: NCT06314490).

### Diagnostic Whole Genome Sequencing

Parental consent was obtained for diagnostic trio or proband WGS and follow-up research. Diagnostic WGS was performed at Rady Children’s Institute for Genomic Medicine (San Diego) as described.^72–74^ Genomic DNA was isolated from whole blood using a Qiagen extraction robot. Sequencing libraries were generated using PCR-free methods (Illumina). Paired 101 nt WGS was performed on NovaSeq 6000 or X Plus instruments (Illumina). Read alignment to the reference human genome and variant calling was performed using the DRAGEN Bio-IT Platform (Illumina). SNVs/indels and CNVs were annotated and analyzed using Enterprise (Fabric Genomics). The median genomic coverage was >30x with a minimum of 90% of Mendelian Inheritance in Man-listed gene coding domains fully covered at ≥10x. Human Phenotype Ontology (HPO) terms used during variant interpretation included seizure (HP:0001250) and other clinical features observed in each case. Nucleotide variants were filtered to retain those with allelic balance of 0.3–0.7 and allele frequency <0.5% in the Genome Aggregation Database. Variants were prioritized by GEM and Phevor (Fabric Genomics) and classified according to American College of Medical Genetics and Genomics and the Association for Molecular Pathology guidelines.^75^ In addition, the Clinical Genome Resource Epilepsy Sodium Channel Variant Curation Expert Panel specifications were utilized for classifying *SCN2A* variants ^76^. For solo proband WGS, segregation analysis of *SCN2A* variants was performed on isolated DNA from parental samples by Sanger sequencing, where available.

### Constellation WGS

Constellation WGS was performed at Illumina Inc. (San Diego) as described.^77^ Archived genomic DNA from diagnostic WGS was used. For each sample, 350 ng of genomic DNA was diluted in tagmentation buffer and loaded into a library strip tube. Two additional reagents were added to the reagent cartridge, and the library reagents were loaded onto a flowcell on a NovaSeq X Plus instrument. Using a custom recipe file, the reagents bind transposomes to the flow cell nanowells. Intact double stranded DNA flows over the flow cell lane, where it binds to transposomes and is cleaved (Fig. 2a). Standard cluster generation and 2x151 cycle sequencing-by-synthesis is performed on the instrument. Read alignment to the reference human genome and variant calling was performed using DRAGEN (Illumina). Cleavage on flow cell nanowells results in clusters in nanowells that originated from the same DNA template molecule being near one another on the flow cell surface, enabling probabilistic modelling of whether read pairs from neighboring clusters originated from the same DNA template molecule. These proximal reads are assigned a unique template read tag, indicating their shared origin. Using proximity information, reads from neighboring clusters are reconstructed into an interspersed version of the original DNA template molecule. As shown in Fig. 2a, where each node represents a read pair deriving from a flow cell cluster, and the lines between them indicate predicted connections between them based on a combination of flow cell and genomic proximity. Connections are derived from a proximity model that provides a Phred-scaled quality score describing the probability that two reads have landed with a certain flow cell displacement and within a given genomic distance by chance. The higher the score, the more likely it is that two reads were derived from the same template molecule. Reads derived from the same template molecule also share the same haplotype. Next, HapCUT2 uses both the template read tags and nucleotide variant calls from DRAGEN to accurately phase the reads into haplotypes^78^. This integrated approach enhances haplotype resolution by leveraging both physical linkage and variant information.

### Allele-specific antisense oligonucleotide development

Using standard WGS from the two index *SCN2A*-CND patients, individualized ASOs were designed to promote selective degradation of the *SCN2A* transcript containing the heterozygous causal variant through recruitment of RNase H1 to the RNA-ASO heteroduplex (Kim-McManus et al., submitted). For each patient, over 500 2’-MOE/DNA gapmers were screened. Firstly, candidate allele specific ASOs were screened in a single dose *in vitro* assay in each patient’s iPSC-derived neurons. Their potency and allele-selectivity were evaluated in a dose response studies in the same assay. ASOs meeting criteria for potency and selectivity were assessed for their potential to trigger innate immunity by measuring CCL22 levels in BJAB cells at concentrations up to 8 µM. Next, ASO candidates were evaluated *in silico* and *in vitro* to confirm absence of effects on any similar human genes. ASOs were then assessed for *in vivo* tolerability in an 8-week single intracerebroventricular dose study in mice and an 8-week single intrathecal dose study in rats. These steps identified an optimal ASO for each patient. nL-SCN2A-001 had the sequence TGomComCoAoAmCAATGTAmCAAoGoGGT, where black nucleotides are unmodified deoxyribose; blue nucleotides are 2’ methoxyethyl deoxyribose, unmarked backbone linkages were phosphorothioate, linkages marked with o were normal phosphodiester, and mC was 5-methylcytosine. Table S2 lists the predicted properties of nL-SCN2A-001. It bound selectively to the reference sequence of intron 23 of *SCN2A* at (GRCh38) Chr 2 g.165375116-165375135 which contains rs190030016 (g.165375126A>G). Selectivity for the causal transcript was due to the index patient being heterozygous for rs190030016 with the alternate allele in trans to the reference nL-SCN2A-001 target sequence. nL-SCN2A-002 had the sequence GGoTomCoAoATTGAAAGATAoTomCmCmC. Table S2 lists the predicted properties of nL-SCN2A-002. It bound selectively to the reference sequence of intron 15 of *SCN2A* at Chr 2:165322220-165322241, which contains rs72874313 (g.165322227A>G). Selectivity for the causal transcript was due to the index patient being heterozygous for rs72874313 with the reference allele *in cis* with the nL-SCN2A-002 target sequence. Finally, nL-SCN2A-001 and -002 were evaluated in two 13-week GLP repeat intrathecal dose toxicology studies in rats. In both studies, the ASOs were considered to be well tolerated. Microscopic findings at necropsy were considered nonadverse, and similar to observations in commercial ASOs. The NOAEL (no-observed-adverse-effect level) was 1 mg/dose for both ASOs.

### SCN2A haplotype analysis in 1000 genome project data

The ASOs nL-SCN2A-002 and nL-SCN2A-001 target common variants rs72874313 (*SCN2A* intron 15) and rs1368238 (*SCN2A* intron 23), respectively. Phased data from the 1kGP were employed to characterize the common haplotypes that these variants tag and identify additional haplotype blocks that may be targets for ASO development^33^. The phased, 30x coverage Chr2 VCF covering 3,202 samples was downloaded from 1kGP ftp site.^79^ Tabix was used to extract the coding domain sequence of *SCN2A* (Chr2: 165,295,823-165,389,822), including encompassed introns. The 3’ and 5’ untranslated regions and introns preceding and trailing the coding domain were excluded. In the interest of finding haplotype blocks consisting of common variants, the minor allele frequency requirement for haplotype block detection was set to 15%. Haplotype blocks were detected using PLINKv1.9 with the following command:

> PLINK --blocks --blocks-max-kb 1000 --blocks-min-maf 0.15

The PLINK output was a collection of haplotype blocks, defined as exclusive SNV sets within which at least 95% of the SNVs pairs have high LD, as measured by D’. An in-house script was used to identify the haplotype sequence patterns and their frequencies observed within each of these haplotype blocks. Haplotype blocks spanning at least 5KB were selected for sequence-level analysis. Genotypes were converted to hapsample format using bcftools convert --hapsample, which produces a matrix in which rows are variants and columns are the haplotypes for each sample (thus each pair of columns represents the diplotype of a single sample). For each haplotype block, the matrix was filtered to the informative SNVs and transposed such that the genotypes could be collapsed into a string indicating the sequence of reference and alternate alleles across the SNVs. These allelic patterns defined the individual haplotypes observed within each haplotype block, and the number of homozygous, heterozygous, and total samples with each haplotype were calculated across the 1kGP super populations. Haplotypes observed at least once in 100 or more samples were selected for visualization, and haplotypes observed in at least 800 1kGP samples (25% of the cohort) were prioritized for ASO sequence prediction.

### Selection of allele-specific ASOs containing SNPs based on 1kGP haplotypes

ASOs containing SNPs mapping to common SCN2A haplotypes in 1kGP samples (Table S1) were identified with ASOG using the Oligogen parameters duplex: RNA-2’Ome-RNA, length 20-22 nt, step 1, salt 150 mM, oligo 10nM, Mfold parameters [Na^+]^ 150 mM, [Mg^2+^] 0.1 mM, folding 37^0^C. Relative energy cutoff 10%, BLASTn parametres cutoff 10%, dB GRCh38.p13, reward 1, penalty 3, gapopen 5, gapextend 2, SpliceAI score threshold 1, G-quadruplexes false for ASO and target. Suitable ASOs were selected based on GC content 40 – 60%, T_m_-NN 55 – 70 ^0^C, T_m_-basic 55 – 70 ^0^C, Tm-salt 55 – 70 0C, Lowest target-sense RNA strands successfully silenced/modified by ASO 40 – 100%, Cumulative target-sense RNA strands successfully silenced/modified by ASO 30 – 100%, and BLASTn query max score limited to the *SCN2A* locus on Chr 2.^44^

## Data Availability

All data produced in the present work are contained in the manuscript

## List of Supplementary Materials

Fig. S1 – S6

Table S1 – S3

## Acknowledgments

We thank the patients and families who participated in this study. We thank the members of the clinical laboratory and the bioinformatics group at RCIGM for diagnostic WGS. A Deo lumen, ab amicis auxilium.

## Funding

This research was supported by California Institute for Regenerative Medicine grant CLIN2-15085 (to OKM). n-Lorem and Illumina provided in-kind funding.

## Author contributions

Each author’s contribution(s) to the paper should be listed [we encourage you to follow the CRediT model]. Each CRediT role should have its own line, and there should not be any punctuation in the initials. Olivia Kim-McManus1,2, Pagé Goddard1, Shahad Olsson1, Liana Protopsaltis1, Joseph Gleeson1,2,3, Qing Zhang4, Nafeesa Kahn5, Ali Crawford5, Stephen F. Kingsmore1*

Examples:

Conceptualization: SFK, OKM, JG

Methodology: SO, PG, QZ, NK, AC

Investigation: SFK, PG

Visualization: SFK, PG, AC

Funding acquisition: OKM

Project administration: LP, SO

Supervision: SFK

Writing – original draft: SFK

Writing – review & editing: OKM, PG, SO, LP, JG, QZ, NK, AC

## Competing interests

QZ, NK and AC are employees and shareholders of Illumina Inc. SFK, OKM and JG have filed a patent related to this work. The other authors declare that they have no competing interests.

## Data and materials availability

All data are available in the main text or the supplementary materials.

---

**Table S1: Characteristics of all *SCN2A* haplotypes located within Chromosome 2 haplotype blocks larger than 5KB observed in the 3,202 1kGP samples.** Chr 2 coordinates are on the GRCh38 reference human genome. SNP Number: Number of informative SNPs (minor allele frequency ≥ 15%) defining a haplotype block. Defining SNP Number: Number of informative SNPs defining a specific haplotype. ASO target SNP contributes to haplotype: A SNP targeted by nL-SCN2A-002 or -001 ASO contributes to the specific haplotype and is represented by the alternate allele. Hom: Number of 1kGP subjects homozygous for specific haplotype. Het: Number of 1kGP subjects heterozygous for specific haplotype. AFR, AMR, EAS, EUR, and SAS: African, Admixed American, East Asian, European, and South Asian 1kGP superpopulations, respectively.

RefAltAllelePattern: Allelic pattern for a specific haplotype across all informative SNPs that define a haplotype block (0 = reference allele, 1 = alternate allele).

**Table S2: Characteristics of 190 common *SCN2A* SNPs defining Chromosome 2 haplotype blocks larger than 5KB observed in the 3,202 1kGP samples and list of samples with heterozygosity for each SNP.** The informative SNPs had minor allele frequency > 15% and are expressed with GRCh38 coordinates as chrom:position:ref sequence:alt sequence. Het: Number of 1kGP subjects heterozygous for given variant across all haplotypes. Hom: Number of 1kGP subjects homozygous for either the alternate allele (alt) or reference allele (ref) of the given variant across all haplotypes. AFR, AMR, EAS, EUR, and SAS: African, Admixed American, East Asian, European, and South Asian 1kGP superpopulations, respectively. Hap Block: Haplotype block the given variant is informative for. n_alt Hap: Number of haplotypes that contain the alternate allele for the given variant. n_ref Hap: Number of haplotypes that contain the reference allele for the given variant.

**Table S3:**
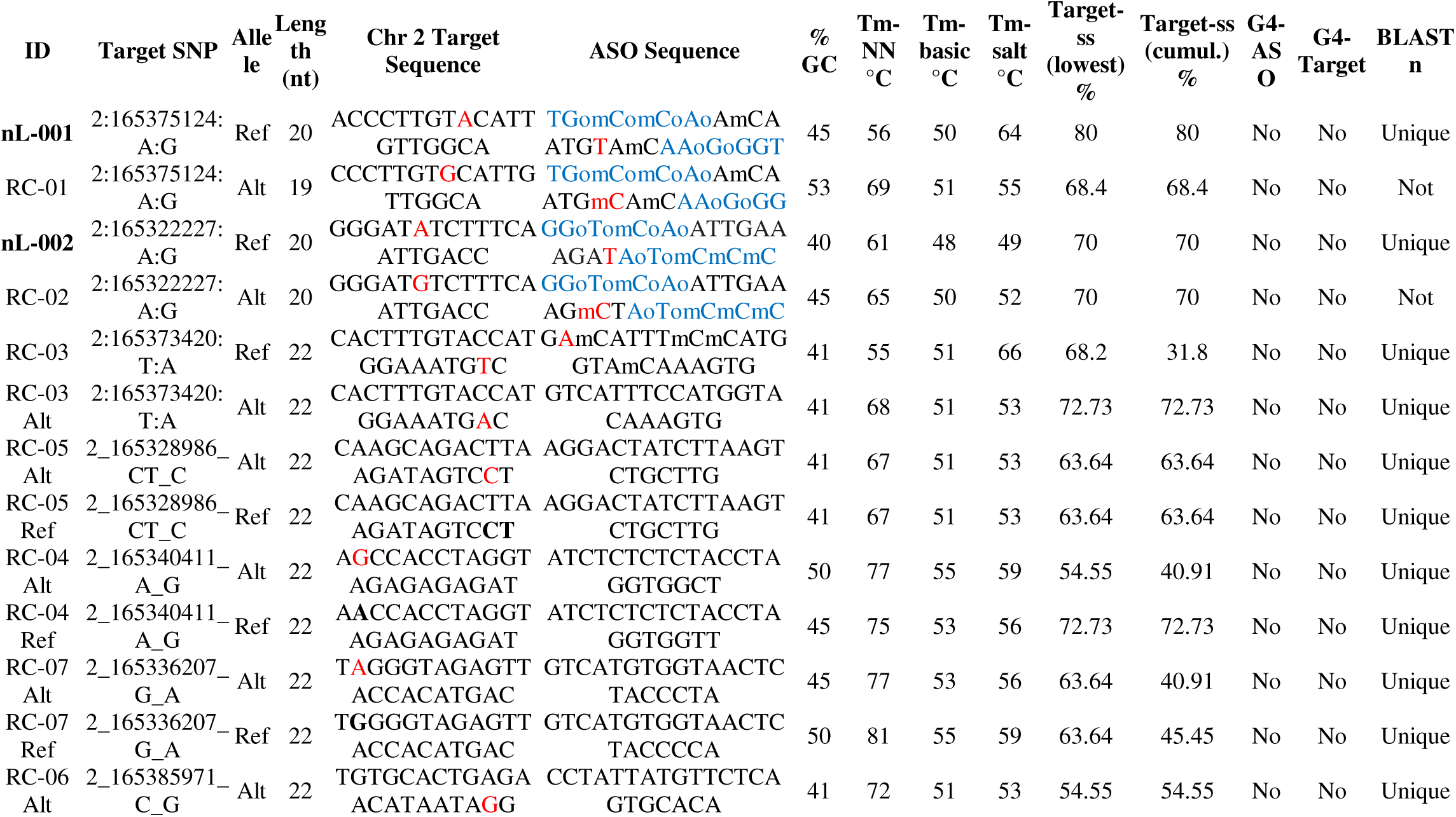
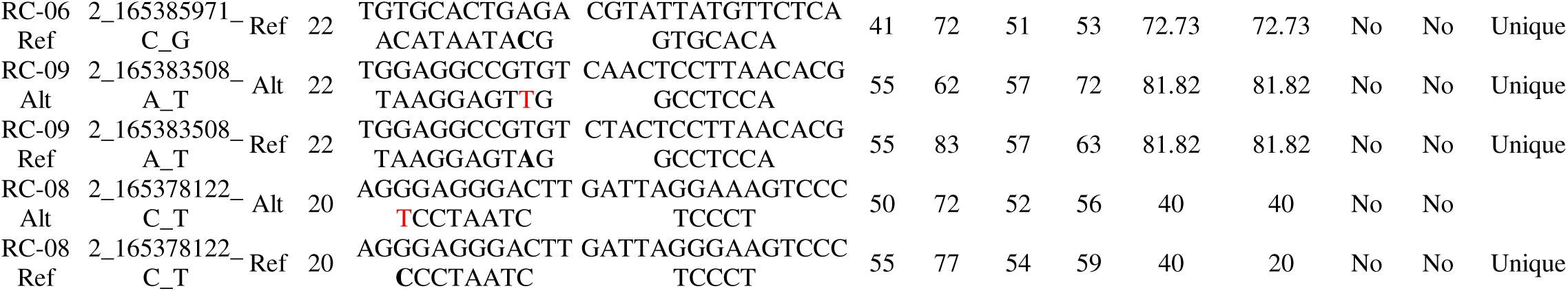
*SCN2A* haplospecific ASO candidate sequences, targets, associated SNPs, and physicochemical properties identified by *in silico* screening. For nL-001, nL-002, RC-01, and RC-02, blue nucleotides are 2’ methoxyethyl deoxyribose, unmarked backbone linkages were phosphorothioate, linkages marked with o were normal phosphodiester, and mC was 5-methylcytosine. Tm-NN, melting temperature using a nearest-neighbor thermodynamic model; Tm-basic, melting temperature using a simple composition-based formula (e.g. Marmur–Doty/“Wallace rule” model); Tm-salt, salt-adjusted Tm correction for 150nM monovalent cation concentration; Target-ss (lowest) %, lowest percentage of target-sense RNA strands being successfully modulated (silenced/modified) by an ASO; Target-ss (cumulative) %, cumulative percentage of target-sense RNA strands being successfully modulated (silenced/modified) by an ASO; G4, guanine nucleotide quadruplex formation.

## Notes

### Clinical Trial

NCT06314490

### Author Declarations

All work was conducted with ethics/IRB approval by the Western Institutional Review Board / Copernicus Group (WCG Clinical) and performed in accordance with the Declaration of Helsinki. N-of-1 clinical trials were approved by the Institutional Review Board at the University of California-San Diego and the Food and Drug Administration.

